# Stress-induced hyperglycemia predicts poor outcomes in primary ICH patients

**DOI:** 10.1101/2024.06.19.24309206

**Authors:** Kevin Gilotra, Jade Basem, Melissa Janssen, Sujith Swarna, Racheed Mani, Benny Ren, Reza Dashti

## Abstract

**Introduction:** The current literature suggests hyperglycemia can predict poor outcomes in patients with primary intracerebral hemorrhage (ICH). Chronic hyperglycemia is seen in patients with pre-existing diabetes (DM), however, acute hyperglycemia in non-diabetic patients is defined as stress-induced hyperglycemia (SIH). This study explored the influence of hyperglycemia on outcomes of primary ICH patients both in the presence and absence of pre-existing DM.

**Methods:** Data regarding admission glucose, pre-existing DM, inpatient mortality, and modified Rankin scale (mRS) scores at discharge were available for 636 patients admitted to Stony Brook Hospital from January 2011 to December 2022 with a primary diagnosis of ICH. Regression models were used to compare outcomes between patients with admission hyperglycemia and/or pre-existing DM to a control group of normoglycemic and non-diabetic ICH patients.

**Results:** Patients with SIH had higher inpatient mortality rates and worse mRS scores at discharge (p<0.001). An association with higher mortality and worse mRS scores at discharge was also seen in patients with hyperglycemia secondary to DM, although the strength of this association was weaker when compared to patients with SIH.

**Conclusion:** In conclusion, our study’s findings suggest that SIH may play a greater role in predicting poor outcomes at discharge rather than a history of poorly controlled DM with chronic hyperglycemia. To develop a more thorough understanding of this topic, prospective studies evaluating the effect of changes in serum glucose during hospital stay on short and long-term outcomes is needed.

## Introduction

Primary intracerebral hemorrhage (ICH) is a devastating condition with a significant rate of mortality and disability (1). ICH forms 10-15% of all strokes, and often requires neurosurgical intervention in addition to comprehensive medical management including strict blood pressure control, and treatment of other underlying medical conditions (2,3). The current literature has shown that a variety of metabolic conditions can contribute to worse outcomes in ICH patients (4). Among them, diabetes mellitus (DM) and stress-induced hyperglycemia (SIH) are believed to be the most common metabolic derangements seen in ICH patients (5). SIH is defined as acute glycemic increase in response to significant injury, irrespective of a pre-existing DM diagnosis (5,6). In the current literature, a glucose level > 180 mg/dL in the setting of an acute stressor such as ICH is the typical cut off for defining SIH in non-diabetic patients (7).

A number of observational studies have demonstrated that SIH is a predictor of mortality, hematoma expansion, and worse functional outcomes amongst primary ICH patients (8–17). Results of INTERACT-2, demonstrated an independent association between both hyperglycemia and DM with 90-day disability and mortality (17). This was further validated in two recent meta-analyses (18,19). When accounting for the presence or absence of DM, majority of studies have shown that SIH predisposes to worse outcomes among ICH patients (14), although others found hyperglycemia in diabetic patients also follows this trend when compared with normoglycemic diabetic patients (9,20). Nevertheless, numerous studies to date have found contradictory outcomes regarding significance of admission SIH, its long-term predictive potential, and its effect when compared to a diabetic population (21–26).

In the present study, our goal was to further explore the association between acute and chronic elevated glucose states with early outcomes in primary ICH. We analyzed patients with primary ICH by categorizing them into different groups based on the presence or absence of previous DM diagnosis and SIH, as defined by admission blood glucose level. We hypothesize that the presence of SIH on admission will be associated with increased disability and mortality.

## Methods

### This study was approved by the Stony Brook University Hospital IRB committee

Medical reports and imaging data was collected retrospectively for all adult patients admitted to Stony Brook University Hospital that presented with intracerebral hemorrhage from January 2011 to December 2022. Inclusion criteria was 1) diagnosis of primary ICH, and 2) age over 18 years old. Exclusion criteria were 1) ICH secondary to underlying vascular lesions, tumors, or trauma 2) hemorrhagic conversion of ischemic stroke 3) missing diagnosis of DM on admission 4) missing blood glucose levels on admission, 5) withdrawal of care or pronouncement of death on admission.

Collected data included basic characteristics, demographics, past medical history, pre-existing medical conditions, laboratory values on arrival and during admission, radiographic findings, and hematoma locations and volume. Modified Rankin Scale (mRS) and Intracerebral Hemorrhage (ICH) scores on admission and discharge were collected as well as social factors such as Do Not Resuscitate/Intubate (DNR/DNI) status and location of discharge. Patients were categorized into four groups based on previous diagnosis of DM and the presence of admission hyperglycemia. SIH was defined as admission glucose >180 mg/dL in non-diabetic patients as in line with previous literature that has used cut-offs ranging from 140-200 for diagnostic purposes (7). *Figure 1* highlights the categorization of patients. The four groups were defined as Group A – (SIH): blood glucose levels on admission above 180 mg/dL without history of DM, Group B – diabetic hyperglycemia (DH): blood glucose levels on admission above 180 mg/dL on admission with previous history of DM, Group C (DM only) – normoglycemia on admission with a history of DM, and Group D (control) – normoglycemia on admission without history of DM. As previously described in the CHEERY study with a similar methodology, (14) the purpose of organizing the groups this way was to determine whether poor outcomes amongst diabetic patients occurred independently of SIH and similarly, if poor outcomes amongst patients with admission hyperglycemia occurred due to SIH or hyperglycemia from underlying DM. For groups A and B, we homogenized our cut-off for both SIH and admission hyperglycemia at a cut-off of 180 mg/dL to compare diabetic patients and non-diabetic patients more effectively with hyperglycemia on admission.

The two primary endpoints were inpatient mortality and level of disability at discharge which was defined by two groups of mRS 0-2 and mRS 3-6 at discharge, with each being considered as “favorable” and “unfavorable” outcome respectively (13). Discharge status to home versus acute care facility or hospice and DNR/DNI status were considered as additional secondary outcome measurements. Figure 2 outlines which binary outcome was considered favorable vs. unfavorable.

For the multivariate regression analysis, Groups A, B, and C were compared to reference Group D as these patients had neither SIH, admission hyperglycemia or DM. We controlled for age, sex and ethnicity for each regression analysis. We used the statistical software R for our analysis of dichotomous and ordinal outcomes. Ordinal logistic regression and standard logistic allow us to compare odds ratios for group effects on the same scale (27–29).

## Results

1006 consecutive patients presented with ICH during this period. Based on exclusion criteria 636 patients with primary ICH, and available glucose levels were included in this retrospective analysis. Table 1 highlights the characteristics of the four groups of patients in this study.

Patients were stratified into one of four groups (Figure 1): non-diabetic patients with SIH (Group A, n=42), diabetic patients with SIH (Group B, n=70), diabetic patients without SIH (Group C, N=101), and non-diabetic patients without SIH (Group D, N=423). Inpatient mortality rate amongst the entire sample was 16.6%. 6.6% of total patients had SIH (mean age = 74, male/female = 25/17), of which 45.2% expired in-hospital. 17.6% of total patients had hyperglycemia of which 36.6% expired in-hospital. 22.9% of total patients had DM of which 21.1% expired in-hospital. When looking within each group, patients with SIH/hyperglycemia with and without DM had nearly double the percentage of mortality compared to the corresponding groups without SIH. Discharge to rehab, acute care facility, or other location was higher than discharge home or with home with hospice for all groups. Group A patients were primarily male (59.5%) while the majority of Group B (and Group C patients were female (57.1% and 58.4%, respectively).

We generated logistic and ordinal logistic regression models that controlled for a variety of pre-existing comorbidities based on what was deemed important in prior studies (14, 27–29), age, sex and ethnicity. Ordinal outcomes were reordered to maintain the same direction of effect for adverse outcomes as standard logistic regression. In our analysis, positive coefficients correspond with an increased probability of adverse clinical outcomes. These included hypertension (HTN), hyperlipidemia (HLD), coronary artery disease (CAD), congestive heart failure (CHF), obesity, prior primary ICH, atrial fibrillation (AF), hematoma location (B and C), malignancy and the presence of anticoagulation or antiplatelet therapy. Table 2 highlights the multivariate logistic and ordinal logistic regression coefficients for Groups A-C to assess their correlation with the primary outcomes (excluding patients without DM and without SIH). A statistically significant association was seen between inpatient mortality and both Group A (p<0.0001) and Group B patients (p=0.0003) but not in the other two groups. Discharge to hospice, acute care facilities and rehabilitation centres was observed significantly more amongst both Group A (p=0.011) and Group B patients (p=0.0016). There was no association between initial hematoma volume and admission glucose, or a history of DM seen across the four groups. When comparing Group A and B, the odds ratios for Group A were higher than what was seen for Group B regarding all primary outcomes including inpatient mortality, unfavorable discharge mRS, and unfavorable discharge status. Group A was noted to have higher mRS scores on admission (p<0.0001) but no association was found with hematoma volume or ICH scores. The remainder of the groups demonstrated no statistically significant association with either hematoma volumes, ICH scores or mRS scores for their presentation on admission. Notably, ORs were also higher for Group A when assessing severity of presentation in comparison to Group B, as demonstrated by worse mRS scores and ICH scores at admission.

## Discussion

Admission hyperglycemia and SIH are both associated with worse outcomes in primary ICH patients (17,19). Furthermore, SIH is thought to be a compensatory response to an acute stressor, such as ICH, in patients without previous diagnosis of diabetes (6). It has been hypothesized that the catecholamine surge in conjunction with many other hormones, particularly cortisol, are responsible for mediating this process by inhibiting glucose uptake into skeletal muscle and increasing gluconeogenesis in the liver (6). Although the purpose of this acute adaptation is to provide the body with more energy in the setting of a physical stressor, acute hyperglycemia can simultaneously cause significant damage to neuronal tissue through oxidative stress and inflammation (6). Other proposed mechanisms for how hyperglycemia leads to neuronal damage include inhibition of cellular proteins such as aquaporin-4 and kallikrein (30,31).

The European Stroke Organization recommends controlling serum glucose through intravenous insulin infusion in a manner that mimics normal physiology while avoiding boluses with high-dose subcutaneous insulin (32). Despite this recommendation, however, some studies have found that tight glycemic control does not necessarily lead to a major improvement in either short-term or long-term outcomes (33,34). In an RCT of 78 patients with SAH, intensive insulin therapy did not lead to significantly improved outcomes (33). A larger meta-analysis of 1248 neurocritical care patients further demonstrated that intensive insulin therapy raises the risk of hypoglycemia and offers minimal mortality benefit (34). Glycemic control in the inpatient setting is often done on a case-by-case basis where the patient’s pre-existing medical history, baseline glucose levels and overall clinical status is collectively taken into consideration. The lack of consensus on how glucose should be monitored in these patients may be attributed to few studies comparing outcomes amongst non-diabetic and diabetic ICH patients with SIH.

To address this management issue, the question now becomes whether poor outcomes in ICH patients are due to the deleterious effects of pre-existing chronic hyperglycemia as seen in poorly controlled diabetics or because of the inflammation, edema and hematoma expansion that occurs in the setting of SIH. While both factors are likely to play a role in these outcomes, the current literature pertaining to this topic is both limited and somewhat conflicting. Godoy et al.’s study of 510 ICH patients demonstrated no significant differences in mortality when comparing those with admission glucose > 160 mg/dL to those with admission glucose < 160 mg/dL (23). On the contrary, Kimura et al. demonstrated that an admission glucose > 150 mg/dL independently predicted inpatient mortality in a sample of 100 ICH patients (8). Meanwhile, another study highlighted that hyperglycemia that persists throughout a hospital stay is associated with higher mortality rates when compared to transient hyperglycemia or normoglycemia (24). The INTERACT-2 study showed that both hyperglycemia and DM were independently associated with long-term disability at 90 day follow-up and higher mortality rates (17).

Few studies have comprehensively compared outcomes between ICH patients with normoglycemia to ICH patients with elevated glucose levels due to SIH or hyperglycemia in the setting of DM. Results from this study showed that patients with SIH (Group A) had a significantly higher risk of unfavorable outcomes when compared to normoglycemic and non-diabetic patients (Group D). Regression coefficients from Table 2 show that Group A patients had a stronger association with inpatient mortality, unfavorable discharge status, and a higher likelihood of being made DNR/DNI compared to all other groups. Both Group A and Group B patients with admission hyperglycemia and DM were significantly more likely to present with higher mRS scores and ICH scores on admission, indicating more severe presentations. Unlike previous studies which set arbitrary cut-offs of hyperglycemia ranging from 140-200 mg/dL, we used the same cut-off of 180 mg/dL to define hyperglycemia in non-diabetic patients, which was deemed as SIH (Group A), and diabetic patients (Group B). Even after homogenizing the cut-off value, our results yielded regression coefficients that demonstrated a much stronger association with unfavorable outcomes amongst ICH patients with SIH (Group A) when compared to regression coefficients for ICH patients with DM and admission hyperglycemia (Group B). In other words, while the combination of DM and admission hyperglycemia was associated with worse outcomes amongst Group B patients, the strength of this association was weaker when compared to the presence of SIH alone in Group A patients. This may support the notion that SIH has a more prominent effect on poor outcomes in ICH patients than hyperglycemia due to DM. Importantly, our study’s findings are very similar to Sun et al.’s study which demonstrated that admission glucose was predictive of poor outcomes at 3-months of follow-up in non-diabetic patients, but admission glucose was not quite predictive in diabetic patients (20).

While poor outcomes associated with SIH may be due to rapid maladaptive pathophysiological changes, chronic hyperglycemia in the setting of DM can also affect outcomes through a more insidious process. Atherosclerotic changes to cerebral microvasculature and endothelial damage can predispose to vessel rupture and subsequently higher volumes of bleeding (14). Perihematomal cell necrosis, cerebral edema and concomitant hematoma expansion in diabetic ICH patients may all serve as plausible mechanisms for why chronic hyperglycemia is associated with poor outcomes (35,36). Additionally, poor outpatient glycemic control may further propagate worse outcomes in diabetic patients (37). Despite this evidence, our study’s results showed no significant associations between unfavorable outcomes amongst patients with a history of DM who had normoglycemia on admission (Group C). This trend may have occurred because Group C patients had relatively well-controlled diabetes, as demonstrated by their normoglycemia on admission. It is important to note that in many prior studies, ICH patients with DM are often compared to those without DM, with little consideration given to serum glucose levels. By comprehensively evaluating serum glucose levels through four individual strata in our study, our results may further support the notion that the deleterious pathophysiology of SIH has a greater effect on outcomes amongst ICH patients than the chronic vasculopathy that occurs due to DM.

Notable strengths of this study include a relatively large sample size amongst each of the individual four strata. This allowed us to perform a complex regression model that compared four groups in an analysis that has rarely been done in the current literature (14). In our regression analysis, we controlled for the presence of multiple CVD risk factors which may have served as confounders of poor outcomes in ICH patients in previous studies.

## Limitations

Our study has a few limitations. Firstly, the retrospective nature of the study limits our ability to determine a direct cause-and-effect relationship between hyperglycemia and poor outcomes. Additionally, this was a single-center analysis subject to institution-dependent biases and it did not assess long-term follow-up after patients had been discharged from the inpatient setting. Another limitation was that pre-admission blood glucose levels and baseline HBA1C values were not available, thus making it difficult to discern the patient’s baseline glycemic control and potential chronic hyperglycemia prior to admission. Similarly, we could not obtain the duration of pre-existing DM in patient-years or distinguish between type one or type two DM. Finally regarding our statistical analysis, because ordinal regression is widely used in the literature (38), we leave further extensions beyond standard ordinal regression to future work.

## Conclusion

Results of this study demonstrate that SIH predicts poor outcomes in ICH patients regardless of their history of DM, although patients without DM had a stronger association with poor outcomes. Glycemic control for non-diabetic patients with SIH greatly differs from those with DM and pre-existing chronic hyperglycemia. SIH patients are given insulin boluses to keep their glucose levels below 180 mg/dL while diabetic patients need additional insulin on top of their basal-bolus regimen and regular diabetic medications. To develop a thorough understanding of the effects of DM and SIH in the role of ICH, larger randomized controlled trials evaluating changes in serum glucose during hospital stay in response to different forms of medical management, and then evaluating long-term outcomes, are needed.

## Data Availability

All data referenced in this manuscript are available upon request from the corresponding author.

## Abbreviations

ICH: intracerebral hemorrhage
SIH: stress-induced hyperglycemia
DM: diabetes mellitus
mRS: modified Rankin Scale
HTN: hypertension
HLD: hyperlipidemia
CAD: coronary artery disease
CHF: congestive heart failure
AF: atrial fibrillation

## Acknowledgements

We would like to acknowledge all members of the Dashti Lab who contributed to the research process for this study.

## Appendices

**Figure 1:**
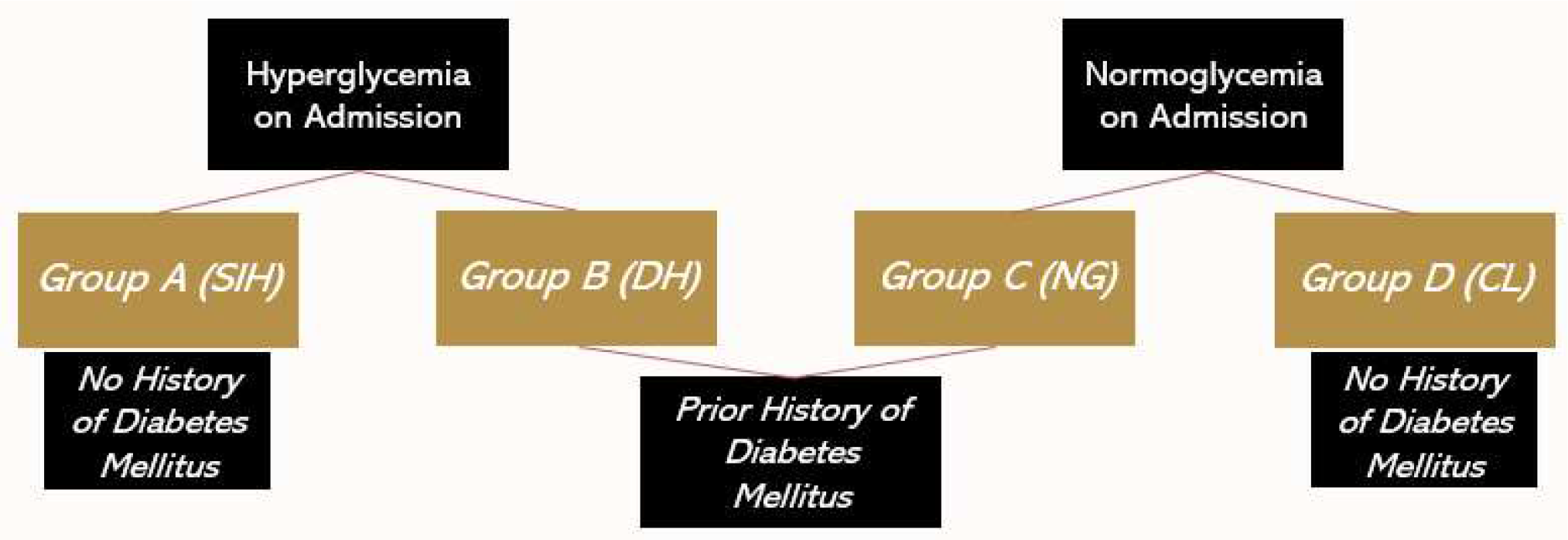
Groups A through D for study design with presence or absence of hyperglycemia on admission and DM indicated. *SIH – stress-induced hyperglycemia, DH – diabetic hyperglycemia, NG – normoglycemia, CL - control

**Figure 2:**
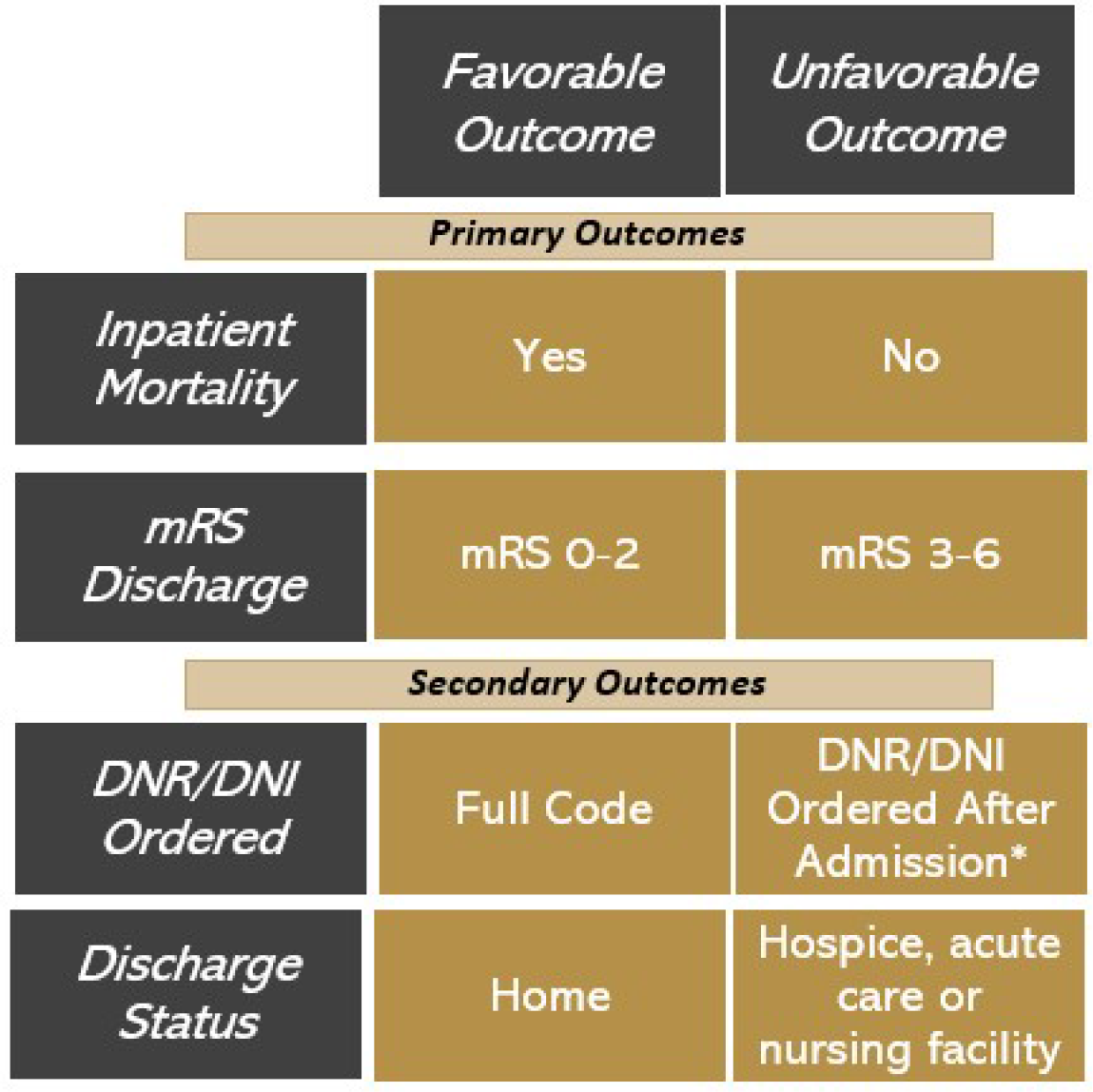
Binary classification of primary outcomes being compared between two groups. *Patients who were already DNR/DNI before admission were excluded from this group.

**Table 1.** ^a^highlights demographic characteristics and clinical outcomes for patients in all four groups.

| **Demographic Information (N=636)<sup>a</sup>** | **Group A - SIH (N=42)** | **Group B – Admission hyperglycemia and history of DM (N=70)** | **Group C – History of DM and normoglycemia on admission (N=101)** | **Group D – Normoglycemia on admission without history of DM (N=423)** |
| --- | --- | --- | --- | --- |
| Median Age | 74 | 71 | 75 | 74 |
| Sex |  |  |  |  |
| Female | 17 (40.5%) | 40 (57.1%) | 59 (58.4%) | 214 (50.6%) |
| Male | 25 (59.5%) | 30 (42.9%) | 42 (41.6%) | 209 (49.4%) |
| Race/Ethnicity |  |  |  |  |
| African American | 0 (0%) | 8 (11.4%) | 10 (9.9%) | 33 (7.8%) |
| Asian | 0 (0%) | 1 (1.4%) | 7 (6.9%) | 6 (1.4%) |
| Hispanic | 0 (0%) | 3 (4.3%) | 3 (3.0%) | 7 (1.7%) |
| White | 42 (100%) | 58 (82.9%) | 81 (80.2%) | 377 (89.1%) |
| **Clinical Outcomes (N=636)** |  |  |  |  |
| Inpatient Mortality | 19 (45.2%) | 22 (31.4%) | 14 (13.9%) | 51 (12.1%) |
| Discharged Home | 1 (2.4%) | 4 (5.7%) | 20 (19.8%) | 103 (24.3%) |
| Discharged to Rehab or Acute Care Facility or Home with Hospice or other | 29 (69.0%) | 43 (61.4%) | 67 (66.3%) | 270 (63.8%) |
| mRS discharge 0-2 | 0 (0.0%) | 7 (10.0%) | 20 (19.8%) | 98 (23.2%) |
| mRS discharge 3 – 6 | 42 (100%) | 63 (90.0%) | 81 (80.2%) | 323 (76.4%) |
| DNR/DNI before admission or within 24 hours | 17 (40.5%) | 9 (12.9%) | 20 (19.8%) | 81 (19.1%) |
| DNR/DNI after 24 hours | 8 (19.0%) | 17 (24.3%) | 11 (10.9%) | 42 (9.9%) |

<sup>a</sup>**Table 1** highlights demographic characteristics and clinical outcomes for patients in all four groups.

**Table 2.** ^a^highlights multivariate logistic and ordinal logistic regression coefficients for Groups A-C (Group D excluded as healthy reference group). Odds ratios, 95% CI and p-values included after controlling for covariates.

| Outcome | Group A<br>– SIH<br>(95% CI) | P-value | Group B –<br>History of<br>DM and<br>normoglycemia on<br>admission<br>(95% CI) | P-value | Group C –<br>Normoglycemia on<br>admission<br>without<br>history of DM<br>(95% CI) | P-value |
| --- | --- | --- | --- | --- | --- | --- |
| Inpatient Mortality | 1.730<br>(0.995,<br>2.465) | <0.001 | 1.169 (0.537,<br>1.801) | <0.001 | -0.110 (-0.801,<br>0.580) | 0.754 |
| Unfavorable mRS discharge (3-6) | N/A* | N/A* | 1.070 (0.171,<br>1.969) | 0.020 | 0.221 (-0.451,<br>0.893) | 0.520 |
| Unfavorable DNR/DNI Status | 1.010<br>(0.299,<br>1.722) | 0.005 | 1.090 (0.500,<br>1.680) | <0.001 | -0.110 (-0.679,<br>0.459) | 0.704 |
| Unfavorable discharge status (hospice, acute care, or nursing facility) | 2.390<br>(0.339,<br>4.441) | 0.022 | 1.785 (0.690,<br>2.879) | 0.001 | 0.283 (-0.354,<br>0.921) | 0.384 |
| Baseline mRS | -0.079 (-0.872,<br>0.714) | 0.845 | -0.231 (-0.833, 0.372) | 0.453 | 0.299 (-0.180,<br>0.778) | 0.222 |
| Admission mRS | 1.775<br>(0.939,<br>2.611) | <0.001 | 0.844 (0.317,<br>1.371) | 0.002 | -0.009 (-0.443,<br>0.424) | 0.966 |
| ICH score | 1.345<br>(0.756,<br>1.934) | <0.001 | 0.682 (0.211,<br>1.154) | 0.005 | -0.260 (-0.680,<br>0.161) | 0.227 |

\*Note that the Group A was not included in the regression due to complete separation, where Group A only consisted of cases in Table 1.

**Figures 3A-3C:**
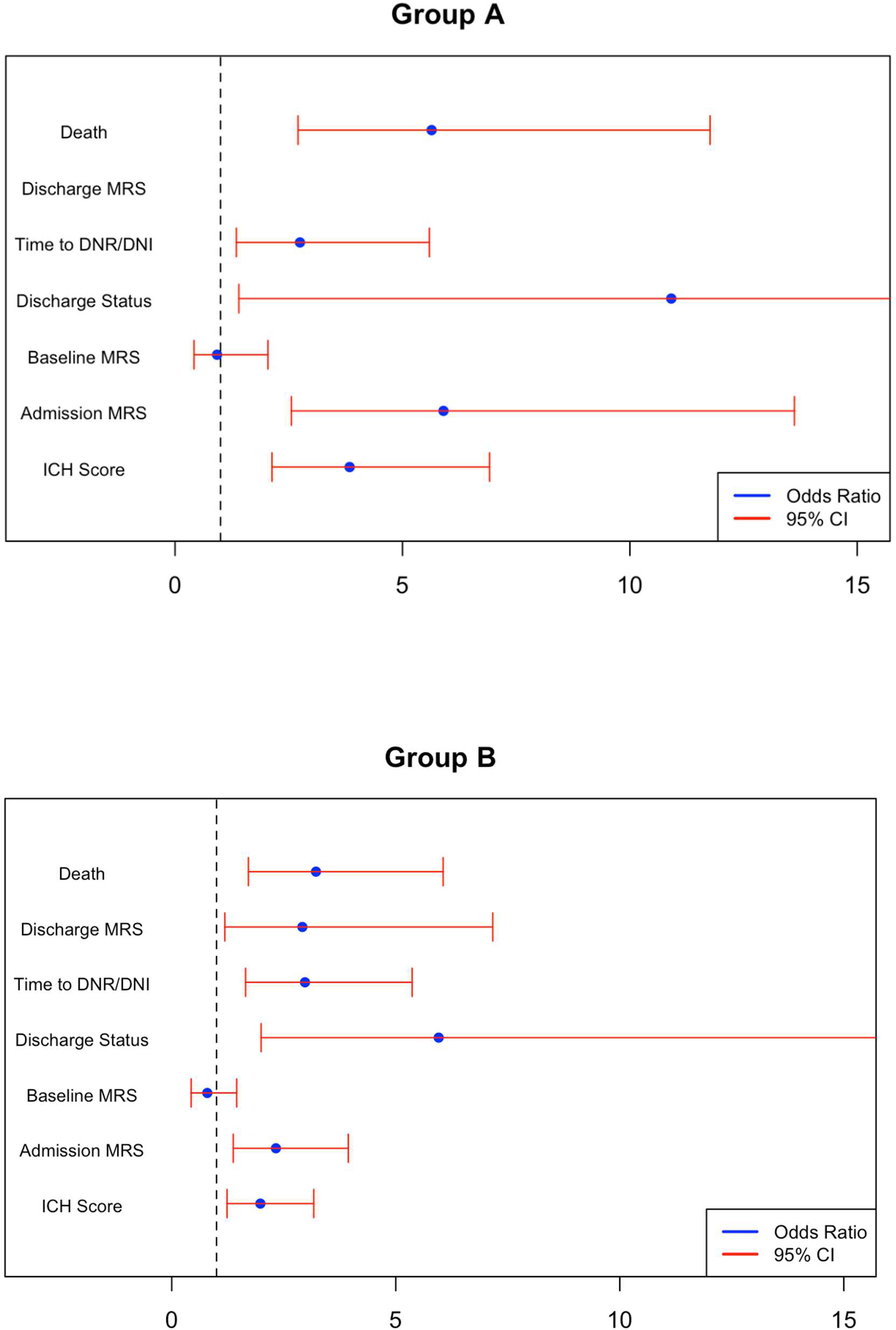
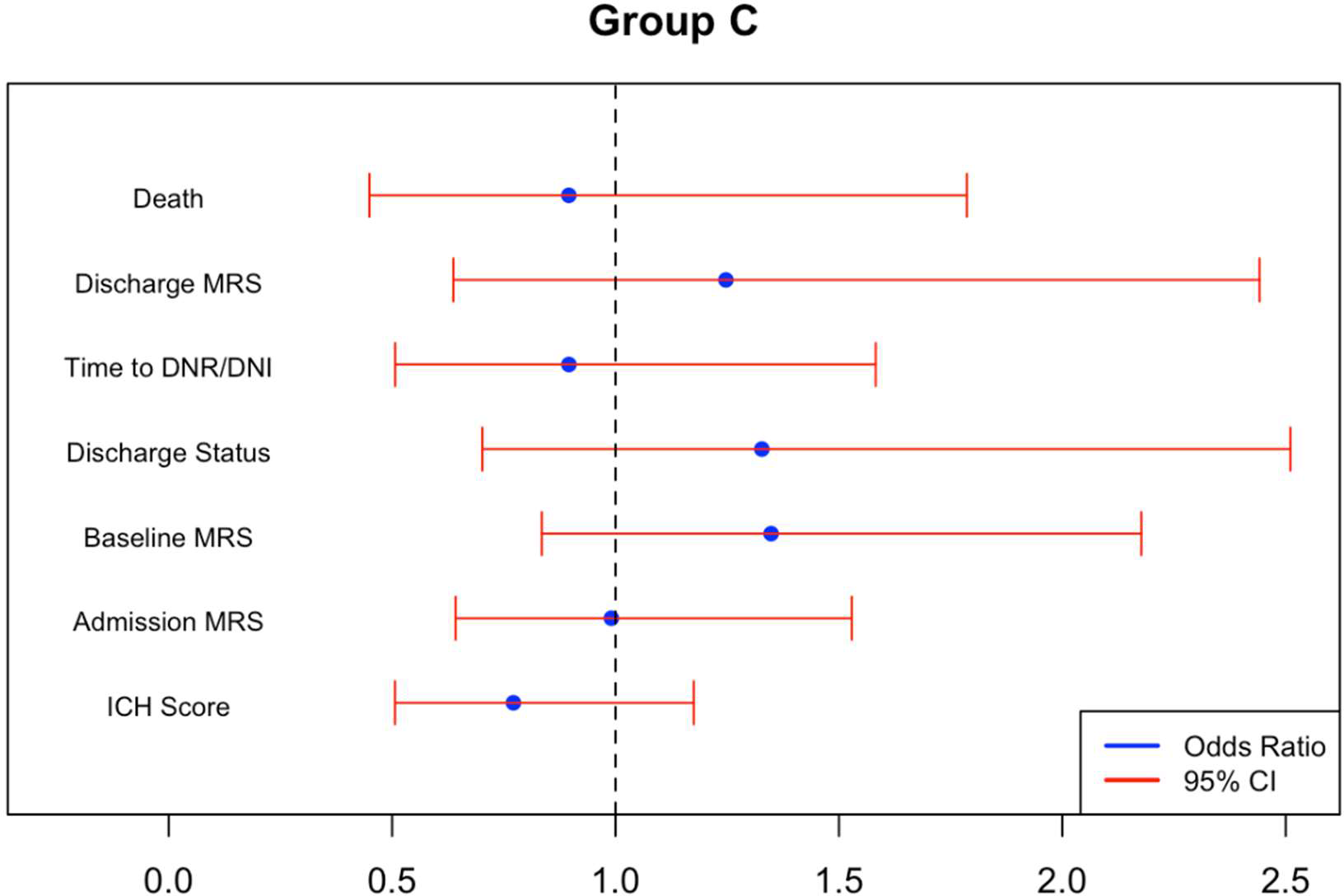
Multivariate ordinal and logistic regression with odds ratios and 95% CI for Group A patients (Figure 3A: SIH), Group B patients (Figure 3B: DM and hyperglycemia on admission) and Group C patients (Figure 3C: normoglycemia on admission and pre-existing DM). All regression models adjusted for history of HTN, HLD, CAD, CHF, obesity, prior ICH, AF, malignancy, anticoagulation and antiplatelet therapy. Odds ratio, OR=1 is plotted as the vertical dashed line with any OR > 1 being considered as an unfavorable outcome.

## References

1. Van Asch CJ, Luitse MJ, Rinkel GJ, Van Der Tweel I, Algra A, Klijn CJ. Incidence, case fatality, and functional outcome of intracerebral haemorrhage over time, according to age, sex, and ethnic origin: a systematic review and meta-analysis. Lancet Neurol. 2010 Feb;9(2):167–76.

2. Stein M, Misselwitz B, Hamann GF, Scharbrodt W, Schummer DI, Oertel MF. Intracerebral Hemorrhage in the Very Old: Future Demographic Trends of an Aging Population. Stroke. 2012 Apr;43(4):1126–8.

3. Sacco S, Marini C, Toni D, Olivieri L, Carolei A. Incidence and 10-Year Survival of Intracerebral Hemorrhage in a Population-Based Registry. Stroke. 2009 Feb;40(2):394–9.

4. Tsao CW, Aday AW, Almarzooq ZI, Anderson CAM, Arora P, Avery CL, et al. Heart Disease and Stroke Statistics—2023 Update: A Report From the American Heart Association. Circulation [Internet]. 2023 Feb 21 [cited 2024 Apr 19];147(8). Available from: https://www.ahajournals.org/doi/10.1161/CIR.0000000000001123

5. McCowen KC, Malhotra A, Bistrian BR. STRESS-INDUCED HYPERGLYCEMIA.

6. Dungan KM, Braithwaite SS, Preiser JC. Stress hyperglycaemia. The Lancet. 2009 May;373(9677):1798–807.

7. Vedantam D, Poman DS, Motwani L, Asif N, Patel A, Anne KK. Stress-Induced Hyperglycemia: Consequences and Management. Cureus [Internet]. 2022 Jul 10 [cited 2024 Apr 19]; Available from: https://www.cureus.com/articles/100945-stress-induced-hyperglycemia-consequences-and-management

8. Kimura K, Iguchi Y, Inoue T, Shibazaki K, Matsumoto N, Kobayashi K, et al. Hyperglycemia independently increases the risk of early death in acute spontaneous intracerebral hemorrhage. J Neurol Sci. 2007 Apr;255(1–2):90–4.

9. Stöllberger C, Exner I, Finsterer J, Slany J, Steger C. Stroke in diabetic and non-diabetic patients: Course and prognostic value of admission serum glucose. Ann Med. 2005 Jan;37(5):357–64.

10. Fogelholm R. Admission blood glucose and short term survival in primary intracerebral haemorrhage: a population based study. J Neurol Neurosurg Psychiatry. 2005 Mar 1;76(3):349–53.

11. Gong Y, Wang Y, Chen D, Teng Y, Xu F, Yang P. Predictive value of hyperglycemia on prognosis in spontaneous intracerebral hemorrhage patients. Heliyon. 2023 Mar;9(3):e14290.

12. Chu H, Huang C, Tang Y, Dong Q, Guo Q. The stress hyperglycemia ratio predicts early hematoma expansion and poor outcomes in patients with spontaneous intracerebral hemorrhage. Ther Adv Neurol Disord. 2022 Jan;15:175628642110706.

13. Li S, Wang Y, Wang W, Zhang Q, Wang A, Zhao X. Stress hyperglycemia is predictive of clinical outcomes in patients with spontaneous intracerebral hemorrhage. BMC Neurol. 2022 Dec;22(1):236.

14. Chen S, Wan Y, Guo H, Shen J, Li M, Xia Y, et al. Diabetic and STRESS-INDUCED hyperglycemia in spontaneous intracerebral hemorrhage: A multicenter prospective cohort (CHEERY) study. CNS Neurosci Ther. 2023 Apr;29(4):979–87.

15. Zhou Y, Luo Z, Yu M, Zhan C, Xu H, Lin R, et al. Acute Phase Blood Glucose Levels and Functional Outcomes in Patients with Spontaneous Intracerebral Hemorrhage. Neuropsychiatr Dis Treat. 2023 Dec;Volume 19:2697–707.

16. Li G, Wang S, Xiong Y, Gu H, Jiang Y, Yang X, et al. Higher fasting blood glucose was associated with worse in-hospital clinical outcomes in patients with primary intracerebral hemorrhage: From a large-scale nationwide longitudinal registry. CNS Neurosci Ther. 2022 Dec;28(12):2260–7.

17. Saxena A, Anderson CS, Wang X, Sato S, Arima H, Chan E, et al. Prognostic Significance of Hyperglycemia in Acute Intracerebral Hemorrhage: The INTERACT2 Study. Stroke. 2016 Mar;47(3):682–8.

18. Zheng J, Yu Z, Ma L, Guo R, Lin S, You C, et al. Association Between Blood Glucose and Functional Outcome in Intracerebral Hemorrhage: A Systematic Review and Meta-Analysis. World Neurosurg. 2018 Jun;114:e756–65.

19. Guo X, Li H, Zhang Z, Li S, Zhang L, Zhang J, et al. Hyperglycemia and Mortality Risk in Patients with Primary Intracerebral Hemorrhage: A Meta-Analysis. Mol Neurobiol. 2016 May;53(4):2269–75.

20. Sun S, Pan Y, Zhao X, Liu L, Li H, He Y, et al. Prognostic Value of Admission Blood Glucose in Diabetic and Non-diabetic Patients with Intracerebral Hemorrhage. Sci Rep. 2016 Aug 26;6(1):32342.

21. Capes SE, Hunt D, Malmberg K, Pathak P, Gerstein HC. Stress Hyperglycemia and Prognosis of Stroke in Nondiabetic and Diabetic Patients: A Systematic Overview. Stroke. 2001 Oct;32(10):2426– 32.

22. Stead LG, Jain A, Bellolio MF, Odufuye A, Gilmore RM, Rabinstein A, et al. Emergency Department Hyperglycemia as a Predictor of Early Mortality and Worse Functional Outcome after Intracerebral Hemorrhage. Neurocrit Care. 2010 Aug;13(1):67–74.

23. Godoy DA, Piñero GR, Svampa S, Papa F, Di Napoli M. Hyperglycemia and Short-term Outcome in Patients with Spontaneous Intracerebral Hemorrhage. Neurocrit Care. 2008 Oct;9(2):217– 29.

24. Wu TY, Putaala J, Sharma G, Strbian D, Tatlisumak T, Davis SM, et al. Persistent Hyperglycemia Is Associated With Increased Mortality After Intracerebral Hemorrhage. J Am Heart Assoc. 2017 Aug 2;6(8):e005760.

25. Zhao Y, Yang J, Zhao H, Ding Y, Zhou J, Zhang Y. The association between hyperglycemia and the prognosis of acute spontaneous intracerebral hemorrhage. Neurol Res. 2017 Feb 1;39(2):152–7.

26. Kang K, Lu J, Ju Y, Wang W, Shen Y, Wang A, et al. Association of pre- and post-stroke glycemic status with clinical outcome in spontaneous intracerebral hemorrhage. Sci Rep. 2019 Dec 13;9(1):19054.

27. Qureshi AI, Huang W, Lobanova I, Chandrasekaran PN, Hanley DF, Hsu CY, et al. Effect of Moderate and Severe Persistent Hyperglycemia on Outcomes in Patients With Intracerebral Hemorrhage. Stroke. 2022 Apr;53(4):1226–34.

28. Yee TW. The VGAM package for negative binomial regression. Aust N Z J Stat. 2020 Mar;62(1):116–31.

29. Yee TW, Stoklosa J, Huggins RM. The VGAM Package for Capture-Recapture Data Using the Conditional Likelihood. J Stat Softw [Internet]. 2015 [cited 2024 Apr 19];65(5). Available from: http://www.jstatsoft.org/v65/i05/

30. Chiu CD, Chen CCV, Shen CC, Chin LT, Ma HI, Chuang HY, et al. Hyperglycemia Exacerbates Intracerebral Hemorrhage via the Downregulation of Aquaporin-4: Temporal Assessment With Magnetic Resonance Imaging. Stroke. 2013 Jun;44(6):1682–9.

31. Liu J, Gao BB, Clermont AC, Blair P, Chilcote TJ, Sinha S, et al. Hyperglycemia-induced cerebral hematoma expansion is mediated by plasma kallikrein. Nat Med. 2011 Feb;17(2):206–10.

32. Fuentes B, Ntaios G, Putaala J, Thomas B, Turc G, Díez-Tejedor E, et al. European Stroke Organisation (ESO) guidelines on glycaemia management in acute stroke. Eur Stroke J. 2018 Mar;3(1):5–21.

33. Bilotta F, Spinelli A, Giovannini F, Doronzio A, Delfini R, Rosa G. The Effect of Intensive Insulin Therapy on Infection Rate, Vasospasm, Neurologic Outcome, and Mortality in Neurointensive Care Unit After Intracranial Aneurysm Clipping in Patients With Acute Subarachnoid Hemorrhage: A Randomized Prospective Pilot Trial. J Neurosurg Anesthesiol. 2007 Jul;19(3):156–60.

34. Kramer AH, Roberts DJ, Zygun DA. Optimal glycemic control in neurocritical care patients: a systematic review and meta-analysis. Crit Care. 2012;16(5):R203.

35. Song EC, Chu K, Jeong SW, Jung KH, Kim SH, Kim M, et al. Hyperglycemia Exacerbates Brain Edema and Perihematomal Cell Death After Intracerebral Hemorrhage. Stroke. 2003 Sep;34(9):2215– 20.

36. Broderick JP, Diringer MN, Hill MD, Brun NC, Mayer SA, Steiner T, et al. Determinants of Intracerebral Hemorrhage Growth: An Exploratory Analysis. Stroke. 2007 Mar;38(3):1072–5.

37. Saliba W, Barnett-Griness O, Gronich N, Molad J, Naftali J, Rennert G, et al. Association of Diabetes and Glycated Hemoglobin With the Risk of Intracerebral Hemorrhage: A Population-Based Cohort Study. Diabetes Care. 2019 Apr 1;42(4):682–8.

38. Ganesh A, Luengo-Fernandez R, Wharton RM, Rothwell PM. Ordinal vs dichotomous analyses of modified Rankin Scale, 5-year outcome, and cost of stroke. Neurology [Internet]. 2018 Nov 20 [cited 2024 May 27];91(21). Available from: https://www.neurology.org/doi/10.1212/WNL.0000000000006554

